# Increased Circulating miR-155 identifies a subtype of preeclamptic patients

**DOI:** 10.1101/2022.03.23.22272742

**Authors:** Zhiyin Wang, Honglei Duan, Yuan Wang, Dan Liu, Chenrui Cao, Yimin Dai, Guangfeng Zhao, Ning Gu, Yan Zhou, Mingming Zheng, Yali Hu

**Affiliations:** Department of Obstetrics and Gynecology, The Affiliated Drum Tower Hospital of Nanjing University Medical School, Nanjing, China; Center for Reproductive Sciences, Department of Obstetrics, Gynecology, and Reproductive Sciences, University of California San Francisco (UCSF), San Francisco, CA 94143

**Keywords:** preeclampsia, heterogeneous, miR-155, serum, subtype

## Abstract

Preeclampsia is a heterogeneous disorder which affect maternal and fetal outcomes. The current classifications of preeclampsia such as “early-“ and “late-” types, and “mild” and “severe” forms, are too imprecise to delineate the pathophysiology of preeclampsia. Here we reported that roughly one third of preeclampsia patients had high expression of maternal serum miR-155 in the case-control study and longitudinal study. The maternal serum miR-155 increased as early as 11-13^+6^ weeks of gestation. The patients with high serum miR-155 had severer clinical symptoms such as higher blood pressure and urine protein, and more adverse maternal and fetal outcomes. Moreover, these patients could be clustered as one group according to clinical manifestation by t-distributed stochastic neighbor embedding analysis. Therefore, these data suggest that preeclamptic patients with high maternal serum miR-155 could be viewed as a subtype of preeclampsia.

## Introduction

Preeclampsia is a common but poorly understood disorder with adverse maternal and fetal outcomes [1-3]. It is recognized that heterogeneity of preeclampsia hinders our understanding of preeclampsia pathophysiology and prevents us from identifying and developing effective therapy for this disorder [4-8].

Currently, preeclampsia is categorized on the basis of timing of presentation into “early-” and “late-” types, and by clinical severity into “mild” and “severe” forms [9, 10]. However, these classifications are too imprecise to delineate the underlying heterogeneous pathophysiology of preeclampsia [4, 5, 7, 11, 12]. Recently, Benton *et al*. classified preeclampsia into “maternal”, “canonical”, and “immunological” subtypes [4], which is helpful for the understanding the different changes in the placentas of preeclamptic patients, but this classification does not allow the phenotypic subtypes to be identified before delivery due to the lack of representative markers in the peripheral circulation in these patients.

One of the well-studied and widely used biomarker for preeclampsia is placental growth factor (PlGF). It is commonly used as one of biomarkers for the screening and diagnosis of preeclampsia and placental dysfunction [13], and might be used to differentiate a subtype of preeclamptic patients [11]. However, as preeclampsia is a heterogeneous disorder, it remains unclear whether there exist other as yet unexamined biomarkers which may represent different subtypes of preeclampsia and therefore could also be utilized in preeclampsia assessment.

There is growing evidence that microRNAs (miRNAs) may play important roles in the development of preeclampsia [14-17]. The placenta is an organ rich in miRNAs, among which, microRNA-155 (miR-155) has been reported as being involved in preeclampsia [18-20]. In our previous study, miR-155 was found to be upregulated in the placentas from severe preeclamptic patients [21]. We also found that the upregulation of miR-155 is able to inhibit cytotrophoblast invasion and vascular relaxation through a targeted effect on cyclin D1 and endothelial nitric oxide synthase [22-24]. Furthermore, placenta-derived miR-155 can be packed into exosomes which are delivered into the maternal circulation [23].

The aim of the present study was to investigate the expression patterns of miR-155 in the circulation of preeclamptic patients, and to understand what proportion of preeclamptic patients exhibits elevated serum miR-155 at presentation, when the elevation of serum miR-155 begins, and what are the clinical characteristics of the patients with elevated miR-155.

## Materials and Methods

### 1. Study design

#### Definitions of preeclampsia and controls

The diagnostic criteria of preeclampsia were new onset hypertension (systolic blood pressure (SBP) sustained at ≥140 mmHg and/or diastolic blood pressure (DBP) sustained at ≥90 mmHg, or both) complicated with proteinuria and/or organ dysfunction after 20 weeks of gestation and the classifications of mild and severe preeclampsia, or early and late onset pattern were based on the criteria proposed by International Society for the Study of Hypertension in Pregnancy (ISSHP) [10]. Controls were defined as uncomplicated normotensive pregnant women who delivered healthy babies. We excluded the patients with chronic hypertension, gestational hypertension, gestational diabetes mellitus, basic renal or heart disease, autoimmune diseases, evidence of infection and non-preeclamptic preterm birth and fetal growth restriction without preeclampsia. Approval for the study was obtained from Ethics Committee of The Affiliated Drum Tower Hospital of Nanjing University Medical School (2016-113-01) and all subjects signed informed consent.

#### Case-control Study Population

We conducted the case-control study between December 2017 and December 2020, including 525 subjects (350 controls and 175 preeclampsia). Some subjects referred from Grass-roots women’ s health care units for their routine antenatal cares after 20 gestational week (GW), who experienced normotensive uncomplicated pregnancies or developed preeclampsia latter and the others were referred from primary hospitals with documented diagnosis of preeclampsia at presentation.

Among 175 preeclamptic patients in cross-sectional study, 116 were mild cases and 59 severe cases, and 64 patients presented early-onset and 111 presented late-onset. The demographic and clinical characteristics in the case-control study subjects were in Table 1.

**Table 1.** Clinical characteristics of subjects in the case-control study.

| Analyzed items | Controls<br>n=350 | PE<br>n=175 | *p*.value |
| --- | --- | --- | --- |
| Maternal age, y | 29.5 ± 3.8 | 30.2 ± 4.9 | >0.05 |
| Pre-pregnancy BMI, kg/m<sup>2</sup> | 24.6 ± 2.8 | 25.1 ± 3.3 | >0.05 |
| GW at serum collection, wks | 35.9 ± 1.7 | 35.2 ± 2.5 | >0.05 |
| GW at delivery, wks | 39.1 ± 2.3 | 35.8 ± 3.6 | <0.01 |
| IVF, n (%) | 38 (10.9) | 29 (16.6) | >0.05 |
| Multipara, n (%) | 53 (15.1) | 30 (17.1) | >0.05 |
| SBP, mmHg | 114.6 ± 8.9 | 156.2 ± 13.8 | <0.01 |
| DBP, mmHg | 72.3 ± 8.6 | 98.8 ± 9.8 | <0.01 |
| Proteinuria, mg/24h | - | 2675.0 ± 2044.0 | >0.05 |
| Birthweight, g | 3684 ± 443.5 | 2672 ± 824.7 | <0.01 |

GW, gestational week; wks, weeks; BMI, body mass index; IVF, in-vitro fertilization; SBP, systolic blood pressure; DBP, diastolic blood pressure; PE, preeclampsia. The SBP and DBP were described as the highest blood pressure during admission.
Continuous variables are expressed as mean ± SD. Discrete variables are expressed as n (percentages).

#### Longitudinal Study Population

The longitudinal cohort subjects were recruited between January 2017 and March 2020. The subjects attended our antenatal clinic from GW 11-13^+6^ for routine antenatal care and were followed up at GW 19-23^+6^, 30-33^+6^, and 35-38^+6^, and delivery respectively. The inclusion criteria were: age ≥ 18 years, singleton pregnancy with a live fetus, early gestational age determined by ultrasound scan. Exclusion criteria were: women with difficult communication or serious mental illness, GW above 13^+6^, fetal abnormality detected at GW 11-13^+6^, and multiple pregnancy. Maternal characteristics, obstetric and medical history, maternal weight and height, maternal blood pressure (BP) and proteinuria were measured and recorded in Viewpoint 6.0 (GE Healthcare GmbH, Germany) as previous report [25]. Written informed consents were obtained from the subjects. Finally, a total of 137 subjects suffered from preeclampsia and 3492 were normotensive uncomplicated pregnant women and delivered normal babies. The exclusion was shown in Supplementary Figure 1.

Although the recruitment time of both studies overlapped for almost two years, the subjects from case-control study and longitudinal study were independent as the pregnant women in cross-sectional study were referred to our hospital after 20 GW and whereas the subjects in longitudinal cohort were recruited as early as 11-13^+6^ GW, and when the patients in longitudinal cohort developed preeclampsia they were recorded into longitudinal study separated from the case-control group.

Among 137 preeclamptic patients in longitudinal cohort, 81 cases were mild and 56 were severe. Based on the GW of preeclampsia onset, 42 patients presented early-onset and 95 presented late-onset. The demographic and clinical characteristics of the longitudinal study subjects are shown in Table 2.

**Table 2.** Clinical characteristics of subjects in the longitudinal study.

| Analyzed items | Controls<br>n=274 | PE<br>n=137 | *p*.value |
| --- | --- | --- | --- |
| Maternal age, y | 29.9 ± 3.5 | 30.3 ± 3.8 | >0.05 |
| Pre-pregnancy BMI, kg/m<sup>2</sup> | 23.3 ± 2.9 | 23.2 ± 3.4 | >0.05 |
| GW at delivery, wks | 39.0 ± 1.5 | 37.8 ± 1.9 | <0.05 |
| IVF, n (%) | 43 (15.7) | 32 (23.4) | >0.05 |
| Multipara, n (%) | 33 (12.0) | 15 (10.9) | >0.05 |
| SBP, mmHg | 118.0 ± 9.3 | 148.6 ± 12.4 | <0.01 |
| DBP, mmHg | 73.00 ± 8.4 | 93.9 ± 10.0 | <0.01 |
| Proteinuria, mg/24h | - | 2837.0 ± 2205.0 | >0.05 |
| Birthweight, g | 3675 ± 543.4 | 2809 ± 625.6 | <0.05 |

BMI, body mass index; GW, gestational week; IVF, in-vitro fertilization; SBP, systolic blood pressure; DBP, diastolic blood pressure; PE, preeclampsia. The SBP and DBP were described as the highest blood pressure during admission.
Continuous variables are expressed as mean ± SD. Discrete variables are expressed as n (percentages).

In the clinical management of preeclamptic patients, the attending obstetricians and the patients were not aware of the test results of miR-155 and PlGF. Therefore, the clinical treatment was not affected by the levels of serum miR-155 or PlGF. The investigators who measured the levels of miR-155 and PlGF were blinded to the clinical data and management.

### 2. Serum Samples

The serum fraction was obtained according to standard protocols. Briefly, the blood samples were collected into standard Vacutainer tubes (367955, SST™ II Advance Plus Blood Collection Tubes, BD Vacutainer) and processed within 1 h by centrifugation at 3000 × g for 15 min at 4 °C. The supernatants were quickly collected, aliquoted, and stored immediately at -80 °C in the Jiangsu Biobank of Clinical Resource within 2 hours. Before use, sera samples were thawed on ice and centrifuged at 2000 × g for 15 min.

A total of 1563 maternal serum samples were available for the longitudinal analysis. Two blood samples were contributed by all subject, taken at GW 11-13^+6^ and 19-23^+6^, and all normotensive controls and some preeclampsia cases provided the other two blood samples at GW 30-33^+6^ and 35-38^+6^ (Supplementary Table 1). In the case-control study, one sample collected from each case at admission to our hospital for preeclampsia or GW matched normotensive pregnant women for routine prenatal examinations (matched within 5 days to the GW from the preeclamptic cases), and total 525 maternal serum samples were available for analysis.

### 3. Serum miRNA isolation and measurement with real-time quantificational polymerase chain reaction (qPCR)

miRNA was isolated from serum (200μL) using the miRNeasy Serum/Plasma Advanced Kit (217204, Qiagen, Germany) according to the manufacturer’ s protocol. cDNA for analyzing miRNAs was prepared form 10ng of total miRNA using a miRCURY LNA RT Kit (339340, Qiagen, Germany). qPCR was performed with a miRCURY LNA SYBR Green PCR Kit (339347, Qiagen, Germany) according to the manufacturer’ s instructions. The levels of miRNAs were analyzed by miRCURY LNA miRNA PCR Assay and el-miR-39 miRCURY LNA miRNA PCR Assay (339306, Qiagen, Germany). The relative expression of miR-155 was analyzed by the 2^-ΔΔCT^ method, and miR-39 was used for normalization [23].

### 4. Measurement of maternal serum PlGF

The maternal serum PlGF was measured using the Cobas e602 system (Roche Diagnostics, Penzberg, Germany). When analyzing the expression pattern of PlGF in normotensive uncomplicated controls, we described the PlGF as original concentration. When analyzing the difference of PlGF between normotensive uncomplicated controls and preeclampsia cases, we converted the concentration to the multiples of the median (MOM) in the GW matched controls.

### 5. Statistical Analysis

The continuous variables with normal distribution are presented as mean ± SD. The continuous variables with abnormal distribution are presented as median (interquartile range). Student’ s t tests were used for continuous variables conforming to normal distribution. Wilcoxon rank sum tests were used for abnormal continuous variables. Chi-square tests were used for comparison of categorical variables and categorical variables were presented as proportions and counts. Values of *p* < 0.05 were considered as statistical significance.

T-distributed stochastic neighbor embedding (t-SNE) was used to analyze the global relationships among all the patients based on the medical histories, clinical presentations, and pregnant outcomes, and to group the patients based on the levels of miR-155 or PlGF.

## Results

### 1. The differences in circulating miR-155 and PlGF levels between preeclamptic cases and normotensive controls in the case-control study

To investigate the expression changes of circulating miR-155 and PlGF in preeclamptic patients, we conducted a case-control study. The results showed that median miR-155 level in the GW matched normotensive pregnant women was 2.5E-3 (from 3.4E-5 to 4.9E-2). However, in 175 preeclamptic patients, miR-155 level was remarkably elevated (1.4E-2 (4.5E-3-3.4E-2) vs. 2.5E-3 (1.1E-3-8.2E-3), *p*<0.0001) (Figure 1A), although in some cases, serum miR-155 levels overlapped with those in controls, indicating that miR-155 was heterogeneous in preeclampsia. To investigate the proportion of preeclamptic patients with high miR-155 levels, we set the P95 of miR-155 levels (2.6E-2) in controls as a cutoff value and divided the subjects into two groups: one group with high miR-155 levels (equal to or above P95) while the other group with normal miR-155 level (below P95). There were 72 subjects exhibiting high miR-155 levels in total 525 subjects, including 31.4% (55/175) preeclamptic patients and 4.9% (17/350) normotensive uncomplicated controls (*p*<0.01), while the remaining 68.8% (120/175) preeclamptic patients had normal circulating miR-155 level (Figure 1B).

**Figure 1.**
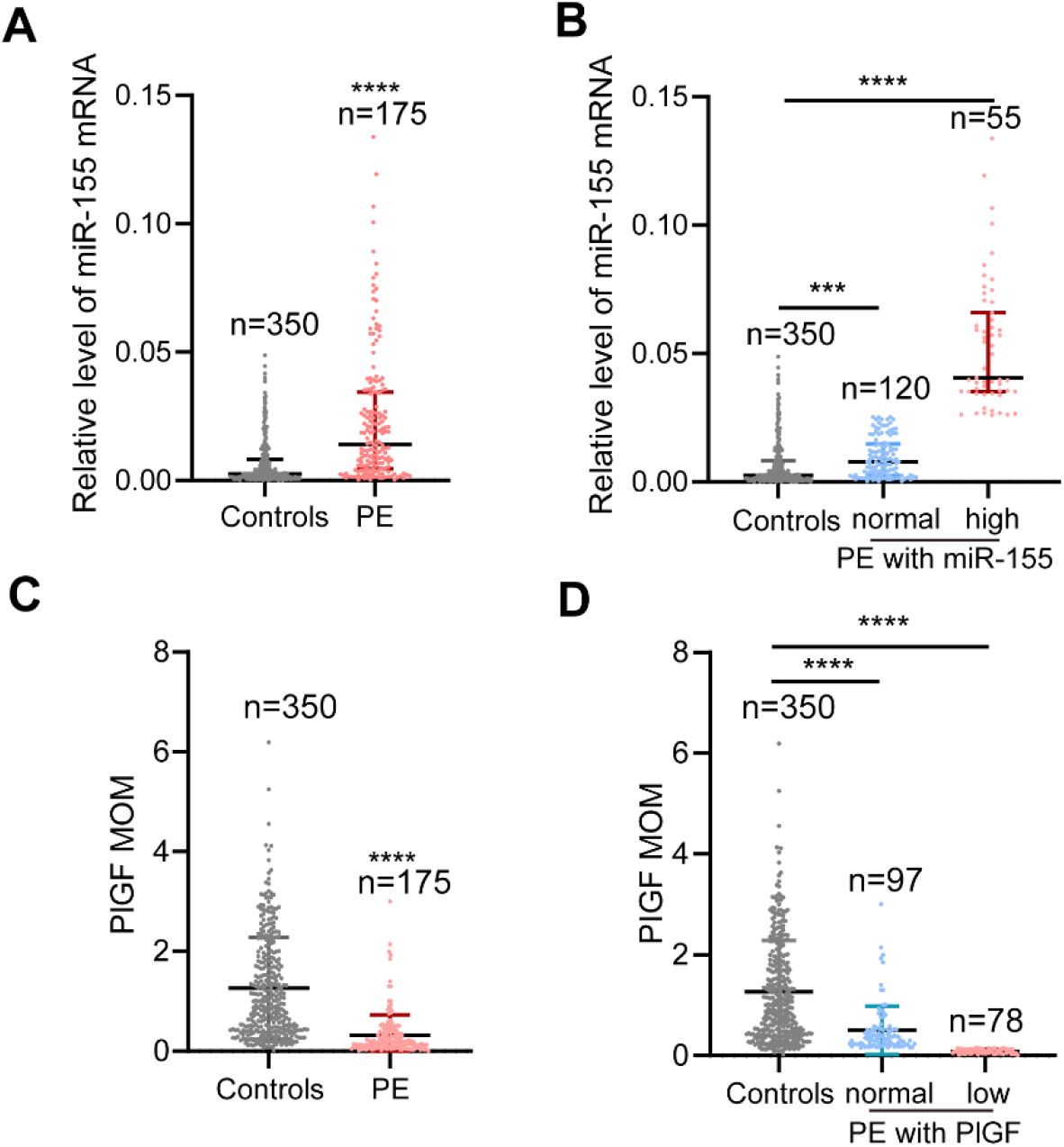
Serum miR-155 increased and PlGF decreased in preeclamptic patients from the case-control study. **A**, Serum miR-155 levels increased in preeclamptic patients compared to controls by qPCR analysis. **B**, Based on the P95 value of controls, there were two groups: one group with high miR-155 levels (equal to or above P95, 8.1 times of controls), as preeclampsia with high miR-155; the other group with miR-155 below P95, as preeclampsia with normal miR-155. **C**, Serum PlGF levels decreased in preeclamptic patients using the Cobas e602 system. **D**, Based on the P5 value of controls, there were two groups: one group with PlGF levels equal to or below P5, as preeclampsia with low PlGF; the other group with PlGF above P5, as preeclampsia with normal PlGF. Data were analyzed with Wilcoxon rank sum tests. Values are median (interquartile range). MOM: multiples of the median. ***, *p*<0.001; ****, *p*<0.0001.

As serum PlGF is an important biomarker for preeclampsia, we also measured the PlGF levels in the subjects. Compared to normal controls, the PlGF levels were significantly low in preeclamptic patients (0.2 (0.1-0.4) vs. 1.0 (0.4-1.9) MOM, *p*<0.0001) but also showed overlap with the controls (Figure 1C), which represented the heterogeneity of PlGF in the preeclampsia. We set P5 of PlGF levels in controls as a cutoff value and divided the subjects into two groups: one showed low PlGF levels (equal to or below the P5) and the other above P5. Of 525 pregnant women, there were 91 subjects exhibited low PlGF levels, including 44.6% (78/175) preeclamptic patients and 4.9% (17/350) normotensive controls (*p*<0.01). In the remaining 55.4% (97/175) preeclamptic cases and 95.1% (333/350) normotensive controls, PlGF levels were above P5 (Figure 1D). Among 78 preeclamptic cases with low PlGF, 24 concurrently had high miR-155 levels. Correlation analysis showed that there is no correlation between miR-155 and PlGF levels in preeclamptic patients (Supplementary Figure 2A).

### 2. Reproducible differences in circulating miR-155 and PIGF levels between preeclampsia and normotensive controls in longitudinal study

To investigate the expressional patterns of serum miR-155 and PlGF throughout the pregnancy and answer when the changes of serum miR-155 and PlGF levels began, we applied the sera of longitudinal cohort to measure the levels of miR-155 and PlGF (controls, n=274; preeclampsia, n=137). The results showed that the median miR-155 levels at GW 11-13^+6^, 19-23^+6^, 30-33^+6^, and 35-38^+6^ were 3.2E-3, 3.3E-3, 3.5E-3, and 3.0E-3, respectively in normotensive uncomplicated controls, and miR-155 levels maintained low across normal pregnancy without significant changes as the progression of pregnancy (Figure 2A). In contrast, the circulating miR-155 levels in preeclamptic patients increased significantly (7.5E-3 (3.1E-3-2.6E-2) vs. 3.2E-3 (1.4E-3-6.4E-3), *p*<0.0001 at GW 11-13^+6^; 6.0E-3 (1.9E-3-2.6E-2) vs. 3.1E-3 (1.0E-3-7.8E-3), *p*<0.0001 at GW 19-23^+6^; 6.7E-3 (1.8E-3-1.9E-2) vs. 3.2E-3 (1.1E-3-7.4E-3), *p*<0.0001 at GW 30-33^+6^; 7.5E-2 (3.1E-3-3.6E-2) vs. 3.0E-3 (1.3E-3-6.7E-3), *p*<0.001 at GW 35-38^+6^respectively) (Figure 2A). However, similar to the case-control study, we also observed that in some patients, serum miR-155 levels overlapped with those in controls during the whole pregnancy. We set the P95 of miR-155 levels in controls as a cutoff value and divided the subjects into two groups: one with high miR-155 levels and the other with normal miR-155 levels (Figure 2B). At GW 11-13^+6^, there were 54 subjects with high miR-155 levels out of a total 411 subjects tested, of whom, 41 cases developed preeclampsia later and they accounted for 75.9% (41/54) in 54 subjects with high miR-155 levels and for 29.9% (41/137) in 137 preeclamptic cases. However, in 274 GW matched normotensive controls, the subjects with high miR-155 levels accounted for 4.7% (13/274). The difference in the rate between preeclamptic cases later and controls with high miR-155 showed statistical significance (*p*<0.05) and the similar incidences and differences of high miR-155 were also observed in GW 19-23^+6^, 30-33^+6^ and 35-38^+6^ between preeclamptic cases and controls (Supplementary Table 2).

**Figure 2.**
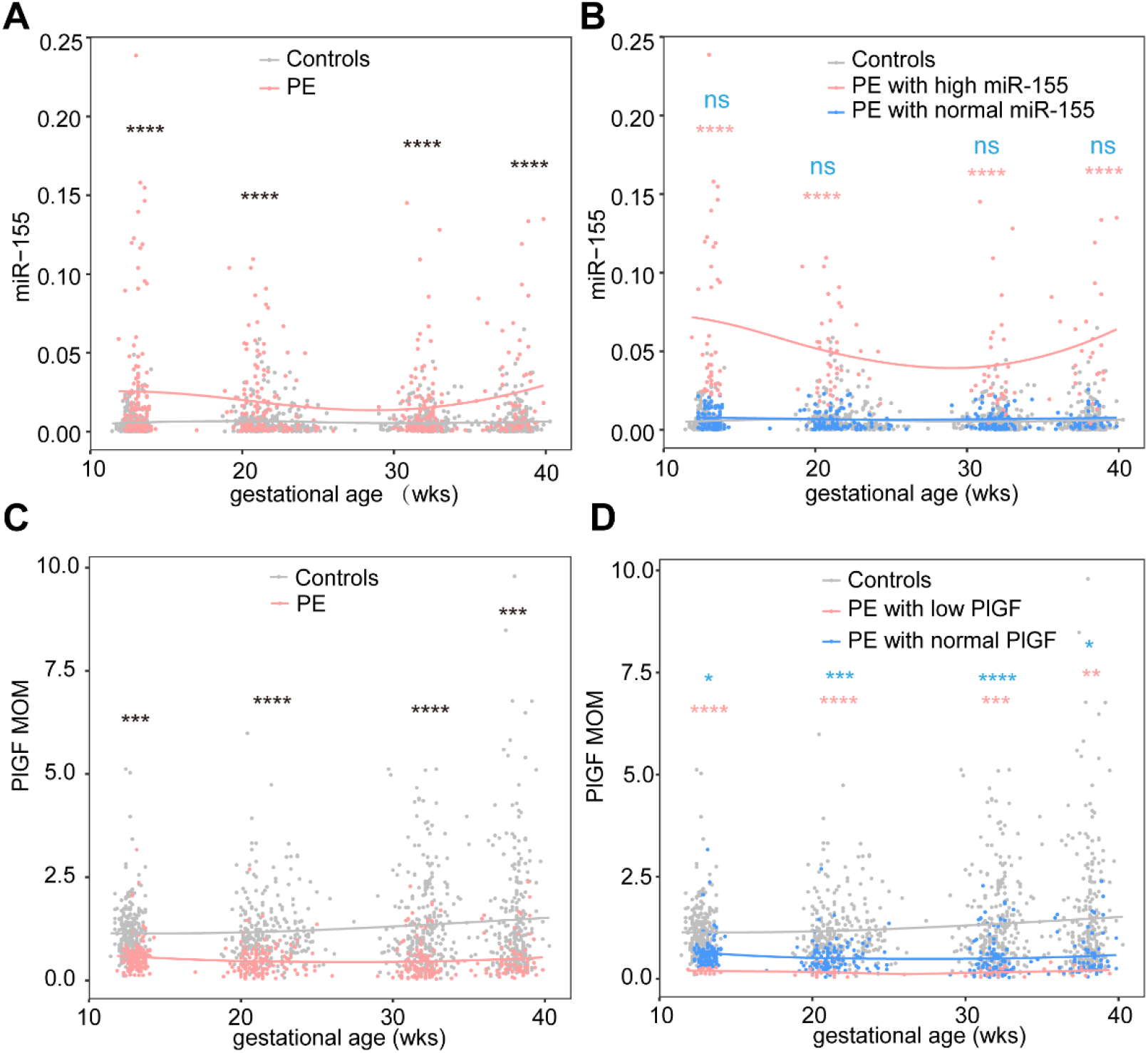
Serum miR-155 increased and PlGF decreased in preeclamptic patients from the longitudinal study. **A**, Serum miR-155 levels in preeclamptic patients were higher than those in controls from GW 11-13^+6^, 19-23^+6^, 30-33^+6^ and 35-38^+6^ respectively. **B**, Based on the P95 value of controls at GW 11-13^+6^, there were two groups: one had remarkably high miR-155 levels (equal to or above P95, 12.3 times of controls), as preeclampsia with high miR-155; in the other group, the level of miR-155 was below P95, as preeclampsia with normal miR-155 level. **C**, Serum PlGF levels in preeclamptic patients were decreased in GW 11-13^+6^, 19-23^+6^, 30-33^+6^ and 35-38^+6^ compared to those in controls respectively. **D**, Based on the P5 value of controls: one showed remarkably low PlGF levels (equal to or below P5, 0.2 times of controls), named as preeclampsia with low PlGF; the other group showed PlGF level above P5, as preeclampsia with normal PlGF. Data were analyzed with Wilcoxon rank sum tests. The curves were shown by curve fitting. MOM: multiples of the median. ns, no significance; *, *p*<0.05; **, *p*<0.01; ***, *p*<0.001; ****, *p*<0.00001. Controls, n=274; PE, n=137.

In 274 normotensive pregnant women, the median serum PlGF level was 51.5 pg/mL at GW 11-13^+6^, increased obviously at GW 19-23^+6^ (375.2 pg/mL, *p*<0.0001) and to peak at GW 30-33^+6^ (548.3 pg/mL, *p*<0.0001) and then decreased to 226.2 pg/mL at GW 35-38^+6^ (Supplementary Figure 3). However, PlGF levels significantly decreased in preeclamptic patients and overlapped with the GW matched controls (GW 11-13^+6^: 0.5 (0.4-0.7) vs. 1.0 (0.7-1.4) MOM, *p*<0.001; GW 19-23^+6^: 0.4 (0.2-0.6) vs. 1.0 (0.7-1.4) MOM, *p*<0.0001; GW 30-33^+6^: 0.3 (0.1-0.6) vs. 1.0 (0.6-1.8) MOM, *p*<0.0001; GW 35-38^+6^: 0.3 (0.2-0.6) vs. 1.0 (0.5-1.9) MOM, *p*<0.001), which represented the heterogeneity nature of PlGF (Figure 2C). We also set P5 of PlGF levels in controls as a cutoff value and divided the subjects into two groups: one with low PlGF levels and the other with normal PlGF levels (Figure 2D). At GW 11-13^+6^, there were 31 subjects with low PlGF levels in a total of 411 subjects. Of whom, 18 cases developed preeclampsia later and they accounted for 58.1% (18/31) in 31 subjects with low PlGF level and for 13.1% (18/137) in 137 preeclamptic cases. However, in 274 GW matched normotensive controls, the subjects with low PlGF levels accounted for 4.7% (13/214). The difference in the rate between preeclamptic cases later and controls with low PlGF showed statistical significance, which was also observed in GW 19-23^+6^, 30-33^+6^ and 35-38^+6^ between preeclamptic cases and controls (Supplementary Table 3). Among 18 preeclamptic cases with low PlGF, 6 concurrently had high miR-155 levels. Correlation analysis showed that there was no correlation between miR-155 and PlGF levels in preeclamptic patients (Supplementary Figure 2B).

### 3. The characteristics in medical history and clinical signs of preeclamptic patients with high miR-155 levels

We combined the data of case-control control study with the longitudinal study, there were totally 312 preeclamptic patients, of whom, 30.8% (96/312) cases exhibited high circulating miR-155, and 31.1% (97/312) presented low PlGF.

Compared with preeclamptic patients with normal miR-155 levels, the cases with high miR-155 had significantly higher frequency of previous preeclampsia history in multiparas (80.0% vs. 73.3%, *p*<0.01), while the cases with low PlGF had less previous preeclampsia history than the patients with normal PlGF levels (64.7% vs. 82.1%, *p*<0.01) (Table 3 and 4). Regarding the clinical presentations, compared to the patients with normal miR-155 levels, the patients with high miR-155 levels presented higher BP, especially in SBP (SBP: 161.4 ± 16.6 vs. 149.5 ± 10.7 mmHg, *p*<0.01; DBP: 97.0 ± 12.5 vs. 94.4 ± 9.3 mmHg, *p*<0.05) and among these patients, the risk of remarkably elevated BP (SBP ≥160 mmHg and/or DBP ≥110 mmHg) was also significantly increased (63.5% vs. 25.0%, *p*<0.05) (Figure 3A and Table 3), therefore the number of patients who needed emergency antihypertensive therapy (≥ 2 times) was increased in high miR-155 group (20.8% vs. 2.3%, *p*<0.01) (Figure 3B and Table 3). The level of urine protein in high miR-155 group was significantly higher (2851 ± 2471 vs. 2380 ± 1914 mg/24h, *p*<0.05) (Table 3). In addition, the patients with high miR-155 showed significant tendency to develop early-onset preeclampsia, in the patients with normal miR-155 level (52.1% vs. 25.9%, *p*<0.01) (Table 3). Contrastingly, when comparing the clinical manifestations of preeclamptic patients with normal or low PlGF level, such dramatic differences as that between patients with and without high miR-155 level, were not observed (Table 3 and 4).

**Table 3.** The medical history, clinical presentations and pregnant outcomes between patients with high miR-155 and normal miR-155.

| Analyzed items | PE |  | *p*.value |
| --- | --- | --- | --- |
|  | Normal miR-155 | High miR-155 |  |
|  | n=216 | n=96 |  |
| Maternal age,y | 30.5 ± 4.4 | 30.3 ± 4.8 | >0.05 |
| Pre-pregnancy BMI, kg/m<sup>2</sup> | 24.4 ± 3.5 | 24.1 ± 3.2 | >0.05 |
| IVF, n (%) | 41 (19.0) | 20 (20.8) | >0.05 |
| Abortion history, n (%) | 27 (12.5) | 31 (32.3) | <0.05 |
| Multipara, n (%) | 30 (13.9) | 15 (15.6) | <0.01 |
| PE history, n (%) | 22 (73.3) | 12 (80.0) | <0.01 |
| Maternal assessment and diagnosis |  |  |  |
| SBP, mm Hg | 149.5 ± 10.7 | 161.4 ± 16.6 | <0.01 |
| DBP, mm Hg | 94.4 ± 9.3 | 97.0 ± 12.5 | <0.05 |
| SBP ≥ 160 and/or DBP ≥ 110 mm Hg, n (%) | 54 (25.0) | 61 (63.5) | <0.01 |
| Proteinuria, mg/24h | 2380 ± 1914 | 2851 ± 2471 | <0.05 |
| Serum creatinine, μM | 50.4 ± 14.6 | 50.5 ± 9.5 | >0.05 |
| Serum uric acid, μM | 401.3 ± 91.9 | 380.8 ± 88.4 | >0.05 |
| AST, U/L | 21.3 ± 13.2 | 31.1 ± 30.6 | <0.01 |
| ALT, U/L | 19.1 ± 7.9 | 23.8 ± 20.2 | >0.05 |
| Platelet, ×10<sup>9</sup>/L | 178.0 ± 52.0 | 159.2 ± 61.3 | <0.01 |
| Platelet < 100×10<sup>9</sup>/L | 7 (3.2) | 17 (17.7) | <0.01 |
| Requiring urgent antihypertensive therapy |  |  |  |
| (≥2 times), n (%) | 5 (2.3) | 20 (20.8) | <0.01 |
| HELLP, n (%) | 2 (0.9) | 5 (5.2) | <0.01 |
| Visual disturbance, n (%) | 10 (4.6) | 15 (15.6) | <0.01 |
| Hypertensive retinopathy, n (%) | 7 (3.2) | 13 (13.5) | <0.01 |
| Placental abruption, n (%) | 2 (0.9) | 7 (7.3) | <0.01 |
| MICU, n (%) | 0 (0.0) | 3 (3.1) | <0.01 |
| The early-onset PE, n (%) | 56 (25.9) | 50 (52.1) | <0.01 |
| Pregnancy outcome |  |  |  |
| GW at delivery, wks | 37.2 ± 2.8 | 35.5 ± 3.6 | <0.01 |
| Male infant, n (%) | 111 (51.4) | 51 (53.1) | >0.05 |
| Birthweight < 3th, n (%) | 10 (4.6) | 21 (21.9) | <0.01 |
| Apgar ≤ 7 at 1 min, n (%) | 5 (2.3) | 6 (6.3) | >0.05 |
| Apgar ≤ 7 at 5 min, n (%) | 5 (2.3) | 3 (3.1) | >0.05 |
| NICU, n (%) | 11 (5.1) | 23 (24.0) | <0.01 |

PE, preeclampsia; BMI, body mass index; IVF, in-vitro fertilization; SBP, systolic blood pressure; DBP, diastolic blood pressure; AST, aspartate aminotransferase ALT; alanine aminotransferase; MICU, maternal intensive care unit admission; GW, gestational week; NICU, neonatal intensive care unit admission. The SBP and DBP were described as the highest blood pressure during admission.
Continuous variables are expressed as mean ± SD. Discrete variables are expressed as n (percentages).

**Table 4.** The medical history, clinical presentations and pregnant outcomes between patients with low PlGF and normal PlGF.

| Analyzed items | PE |  | *p*.value |
| --- | --- | --- | --- |
|  | Normal PIGF<br>n=215 | Low PIGF<br>n=97 |  |
| Maternal age, y | 30.4 ± 4.2 | 30.55 ± 4.74 | >0.05 |
| Pre-pregnancy BMI, kg/m<sup>2</sup> | 24.2 ± 3.4 | 24.39 ± 3.48 | >0.05 |
| IVF, n (%) | 38 (17.67) | 23 (23.7) | >0.05 |
| Abortion history, n (%) | 40 (18.6) | 18 (18.6) | >0.05 |
| Multipara, n (%) | 28 (13.0) | 17 (17.5) | <0.01 |
| PE history, n (%) | 23 (82.1) | 11 (64.7) | >0.05 |
| Maternal assessment and diagnosis |  |  |  |
| SBP, mm Hg | 150.6 ± 10.1 | 154.9 ± 15.8 | <0.01 |
| DBP, mm Hg | 94.1 ± 9.5 | 96.5 ± 11.5 | <0.05 |
| SBP ≥ 160 and/or DBP ≥ 110 mm Hg, n (%) | 78 (36.3) | 37 (38.1) | >0.05 |
| Proteinuria, mg/24h | 2514 ± 1945 | 2755 ± 2376 | >0.05 |
| Serum creatinine, μM | 50.8 ± 7.9 | 50.1 ± 16.2 | >0.05 |
| Serum uric acid, μM | 393.4 ± 98.8 | 396.1 ± 84.0 | >0.05 |
| AST, U/L | 24.4 ± 17.0 | 24.2 ± 23.0 | >0.05 |
| ALT, U/L | 19.8 ± 12.5 | 23.0 ± 20.9 | >0.05 |
| Platelet, $\times 10^9$/L | 170.1 ± 52.9 | 173.6 ± 57.2 | >0.05 |
| Platelet < 100 $\times 10^9$/L | 17 (7.9) | 7 (7.2) | >0.05 |
| Requiring urgent antihypertensive therapy |  |  |  |
| (≥ 2 times), n (%) | 16 (7.4) | 9 (9.3) | >0.05 |
| HELLP, n (%) | 5 (2.3) | 2 (2.1) | >0.05 |
| Visual disturbance, n (%) | 17 (7.9) | 8 (8.3) | >0.05 |
| Hypertensive retinopathy, n (%) | 15 (7.0) | 5 (5.2) | >0.05 |
| Placental abruption, n (%) | 5 (2.3) | 4 (4.1) | >0.05 |
| Heart failure, n (%) | 2 (0.9) | 0 (0.0) | >0.05 |
| MICU, n (%) | 3 (1.4) | 0 (0.0) | >0.05 |
| The early-onset PE, n (%) | 76 (35.3) | 30 (30.9) | >0.05 |
| Pregnancy outcome |  |  |  |
| GW at delivery, wks | 37.0 ± 2.9 | 36.7 ± 5.3 | >0.05 |
| Male infant, n (%) | 113 (52.6) | 49 (50.5) | >0.05 |
| Birthweight < 3th, n (%) | 16 (7.4) | 15 (15.5) | <0.01 |
| Apgar ≤ 7 at 1 min, n (%) | 7 (3.3) | 4 (4.1) | >0.05 |
| Apgar ≤ 7 at 5 min, n (%) | 4 (1.7) | 4 (4.1) | >0.05 |
| NICU, n (%) | 18 (8.4) | 16 (16.5) | <0.05 |

PE, preeclampsia; BMI, body mass index; IVF, in-vitro fertilization; SBP, systolic
blood pressure; DBP, diastolic blood pressure; AST, aspartate aminotransferase ALT; alanine aminotransferase; MICU, maternal intensive care unit admission; GW, gestational week; NICU, neonatal intensive care unit admission; PlGF, placental growth factor. The SBP and DBP were described as the highest blood pressure during admission. Continuous variables are expressed as mean ± SD. Discrete variables are expressed as n (percentages).

**Figure 3.**
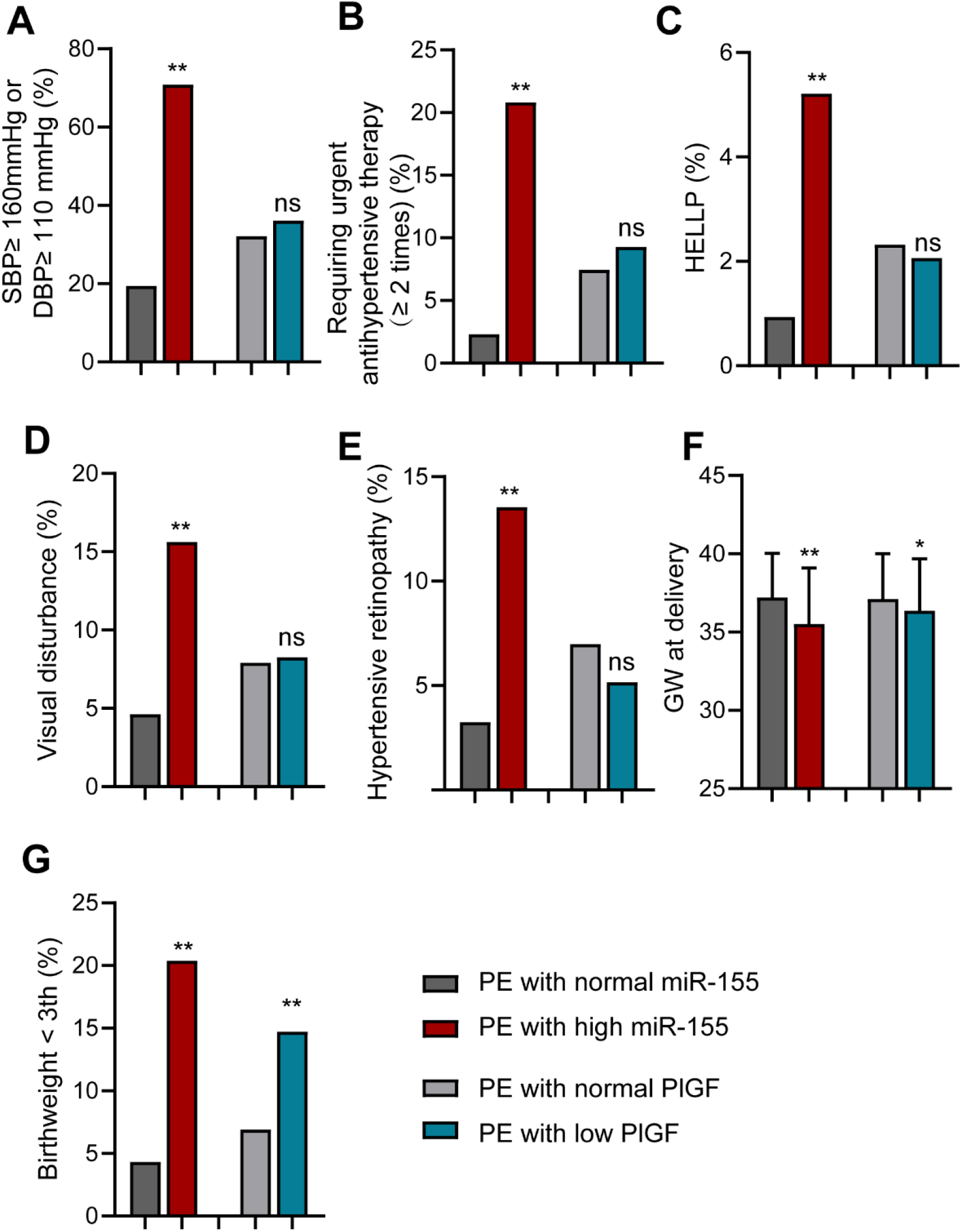
miR-155-RE patients exhibited severer clinical symptoms. **A**, The rate of SBP ≥160 or DBP ≥ 110mmHg in high miR-155 group increased compare to that in normal miR-155 group while there was no significant difference between the cases with low PlGF and normal PlGF. **B**, The rate of requiring urgent antihypertensive therapy (≥ 2 times) in high miR-155 group increased while there was no significant difference between the cases with low PlGF and normal PlGF. **C-E**, The rates of HELLP, visual disturbance and hypertensive retinopathy in high miR-155 group were all higher than those in normal miR-155 group while there were no significant difference between the cases with low PlGF and normal PlGF. **F**, The GWs at delivery were earlier in high miR-155 or low PlGF group than those in normal miR-155 or PlGF group. **G**, The rates of birthweight < 3th percentile in high miR-155 or low PlGF group were higher than those in normal miR-155 or PlGF group. **A-E** and **G** were analyzed with chi-square tests. **F** was analyzed with student’ s t tests. SBP, systolic blood pressure; DBP, diastolic blood pressure; GW, gestational age; ns, no significance; *, *p*<0.05; **, *p*<0.01, ***, *p*<0.001.

### 4. More adverse maternal and fetal outcomes in preeclamptic patients with high miR-155 levels

As to the maternal outcomes, there was no maternal death in this study. The patients admitted to the intensive care unit due to heart failure, pulmonary edema, acute renal failure and eclampsia accounted for 3.1% in high miR-155 group, but none in patients with normal miR-155 level (*p*<0.01) (Table 3). Compared with patients with normal miR-155 levels, the incidences of HELLP syndrome (5.2% vs. 0.9%, *p*<0.01), visual disturbance (15.6% vs. 4.6%, *p*<0.01), hypertensive retinopathy (13.5% vs. 3.2%, *p*<0.01), and placenta abruption (7.3% vs. 0.9%, *p*<0.01) in patients with high miR-155 level were significantly increased (Figure 3C-E and Table 3).

Regarding fetal outcomes, there was no perinatal death in this study. The rate of neonatal intensive care unit admission (NICU) in high miR-155 group was significantly higher than that of the group with normal miR-155 group (24.0% vs. 5.1%, *p*<0.05) (Table 3). The mean GW of termination of pregnancy was lower (35.5 ± 3.6 vs. 37.2 ± 2.8 weeks, *p*<0.01) and the rate of birthweight < 3th percentile was significantly increased (21.9% vs. 4.6%, *p*<0.01) in high miR-155 group (Figure 3F and Table 3). In patients with low PlGF, there were more neonates who required admission in NICU (16.5% vs. 8.4%, *p*<0.05) and who had birthweight < 3th percentile (15.5% vs. 7.4%, *p*<0.01) than those in the group with PlGF above P5 group (Figure 3G and Table 4). However, in normotensive controls with high miR-155 and or with low PlGF, the birthweight was normal, although slightly lower than that of the normotensive controls with normal miR-155 or PlGF (Supplementary Table 4 and 5).

The combination of high miR-155 and low PlGF occurred in 30 preeclamptic patients. Of them, 76.7% (23/30) fulfilled the diagnose of severe preeclampsia, 26.7% (8/30) needed more than twice emergent antihypertensive treatment, and 20.0% (6/30) were complicated with hypertensive retinopathy. There was a propensity that these patients had more abovementioned manifestations than those with high miR-155 level alone but the differences had no statistical significance (Supplementary Table 6). In addition, the rate of birthweight <3th percentile was slightly higher in patients with both high miR-155 and low PlGF than that in the patients with high miR-155 pattern alone (23.3% vs. 21.9%, *p*<0.05) (Supplementary Table 6).

### 5. The preeclamptic patients with high circulating miR-155 level can be clustered as one group

To further investigate the heterogeneity of the biological patterns of high miR-155 and/or low PlGF, we used t-SNE analysis to visualize the high dimensional relationships between the medical history, clinical presentations, pregnant outcomes and the levels of circulating miR-155 or PlGF in 2-dimensional space. The results showed that the patients with high miR-155 pattern were predominantly clustered on the left of the plot, whereas the patients with normal miR-155 level showed dispersed distribution (Figure 4A). On the contrary, the patients with low or high PlGF levels did not show clear clusters (Figure 4B).

**Figure 4.**
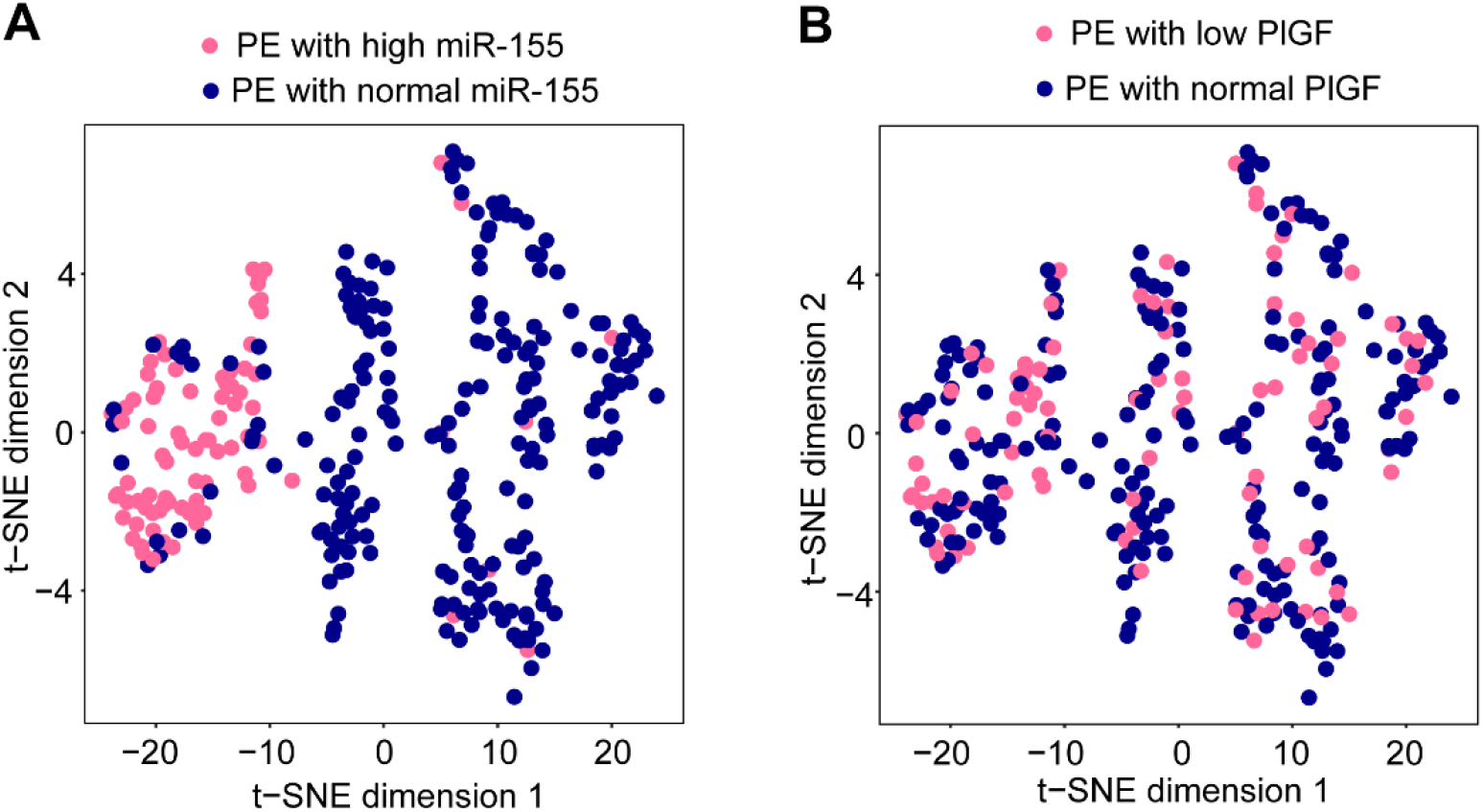
Patients with high miR-155 levels presented a subtype of PE. **A**, Preeclamptic patients with high miR-155 were predominantly clustered on left of the plot as analyzed by t-SNE analysis. B, Both preeclamptic patients with low and normal PlGF could not be clustered by t-SNE analysis.

## Discussion

In this study, we demonstrated that about 30% preeclamptic patients presented remarkable elevation of sero-miR-155 who had more severe clinical manifestations and more adverse maternal and fetal outcomes. And noteworthily, those patients could be well clustered as a group, indicating that the patients with high sero-miR-155 level had relatively homogenous feature. So, measuring circulating miR-155 may be helpful to further differentiate the heterogeneous preeclampsia.

Unlike previous studies [23, 26], in this study, we applied the case-control study and longitudinal study, in which the subjects from both studies were independent. Through the case-control study with a large sample size, we observed that about 30% preeclamptic patients presented high sero-miR-155 level and through longitudinal study we not only found the similar proportion of preeclamptic patients with high sero-miR-155 but also observed that the increase of miR-155 level took place as early as at GW11-13^+6^ and maintaining high level across pregnancy, suggesting that certain proportion of preeclamptic patients with high sero-miR-155 level indeed existed and miR-155 was not fit for an uniform predictor for preeclampsia. However, the patients with high miR-155 level had some clinical characteristics in terms of previous preeclampsia history, and manifestations and based on the features, the patients with high miR-155-level could be well clustered as one group, suggesting that circulating high level of miR-155 was able to represent a subtype in preeclamptic patients and we could take the advantage to identify the subgroup of preeclamptic patients in the first trimester.

PlGF is an angiogenic factor, with known low serum level in placenta-mediated disease especially in preeclampsia [13]. Chappell LC *et al*. reported that in women presenting before 35 weeks’ gestation with suspected preeclampsia, low PlGF has high sensitivity for the diagnosis of preeclampsia within 14 days [27]. Recently, on behalf of the PARROT trial group, Duhig *et al*. reported that in a real world setting the availability of PlGF test results substantially reduces the time to clinical confirmation of preeclampsia [28].

In this study, we also demonstrated that PlGF in preeclampsia was low overall, with some of the patients below the P5 in normal controls and the greatest difference occurred after 20 GW, which was similar to the findings of previous reports [11, 29]. However, although the patients with low PlGF level showed more severe hypertension and more FGR, the t-SNE analysis showed that those patients could not be well clustered as a group (Figure 4B), suggesting that the clinical heterogeneity may exist in the patients with low PlGF level.

## Strengths and limitations

A major strength of this study is our well-defined cohort, including a longitudinal study related to preeclampsia and a preeclampsia cross sectional study and the findings in the cross-sectional study was reproduced in longitudinal study. In addition, the collection of clinical data was comprehensive, the samples processing had good quality and the definition of preeclampsia was standardized.

We demonstrate that circulating miR-155 level had the potential as a biomarker for a subtype of patients with preeclampsia. This is a step forward in the development of stratified medicine in obstetrics where targeted pathogenesis research and therapy could be applied.

However, there were several limitations in the present study. First, this study was performed just in one hospital, and the results were not tested by other centers. Second, the correlation of high miR-155 level with other parameters for classification of preeclampsia remains clarified. Third, the relationship between circulation level of miR-155 and placental level of miR-155 in the subtype of preeclamptic patients with high miR-155 need to be studied since preeclampsia is believed to be triggered by placental factors.

## Data Availability

All data produced in the present work are contained in the manuscript

## Acknowledgements

This work was supported by National Key R&D Program of China (2021YFC2701603); Jiangsu Provincial Key Medical Center (YXZXB2016004); National Natural Science Foundation of China (82071666); Jiangsu Biobank of Clinical Resources (BM2015004).

## Author’ s contributions

Z.Y.W., H.L.D and Y.W.: carried the experiments and drafted the manuscript; D.L., Y.M.D and C.R.C.: helped collecting clinical samples; N.G. and G.F.Z.: analyzed the data and draw graphs; Y.Z., M.M.Z. and Y.L.H.: designed the study and supervisor of this study.

## Conflict of interest

The authors declare no competing interests.

## Notes

### Competing Interest Statement

The authors have declared no competing interest.

### Funding Statement

This study was funded by National Key R&D Program of China (2021YFC2701603); Jiangsu Provincial Key Medical Center (YXZXB2016004); National Natural Science Foundation of China (82071666); Jiangsu Biobank of Clinical Resources (BM2015004).

### Author Declarations

Ethics Committee of The Affiliated Drum Tower Hospital of Nanjing University Medical School gave ethical approval for this work

