## Supplementary for "Increased Circulating miR-155 identifies a subtype of preeclamptic patients"

**Abstract**

Preeclampsia is a heterogeneous disorder which affect maternal and fetal outcomes. The current classifications of preeclampsia such as “early-” and “late-” types, and “mild” and “severe” forms, are too imprecise to delineate the pathophysiology of preeclampsia. Here we reported that roughly one third of preeclampsia patients had high expression of maternal serum miR-155 in the case-control study and longitudinal study. The maternal serum miR-155 increased as early as 11-13^+6^ weeks of gestation. The patients with high serum miR-155 had severer clinical symptoms such as higher blood pressure and urine protein, and more adverse maternal and fetal outcomes. Moreover, these patients could be clustered as one group according to clinical manifestation by t-distributed stochastic neighbor embedding analysis. Therefore, these data suggest that preeclamptic patients with high maternal serum miR-155 could be viewed as a subtype of preeclampsia.

**Key words:** preeclampsia; heterogeneous; miR-155; serum; subtype;

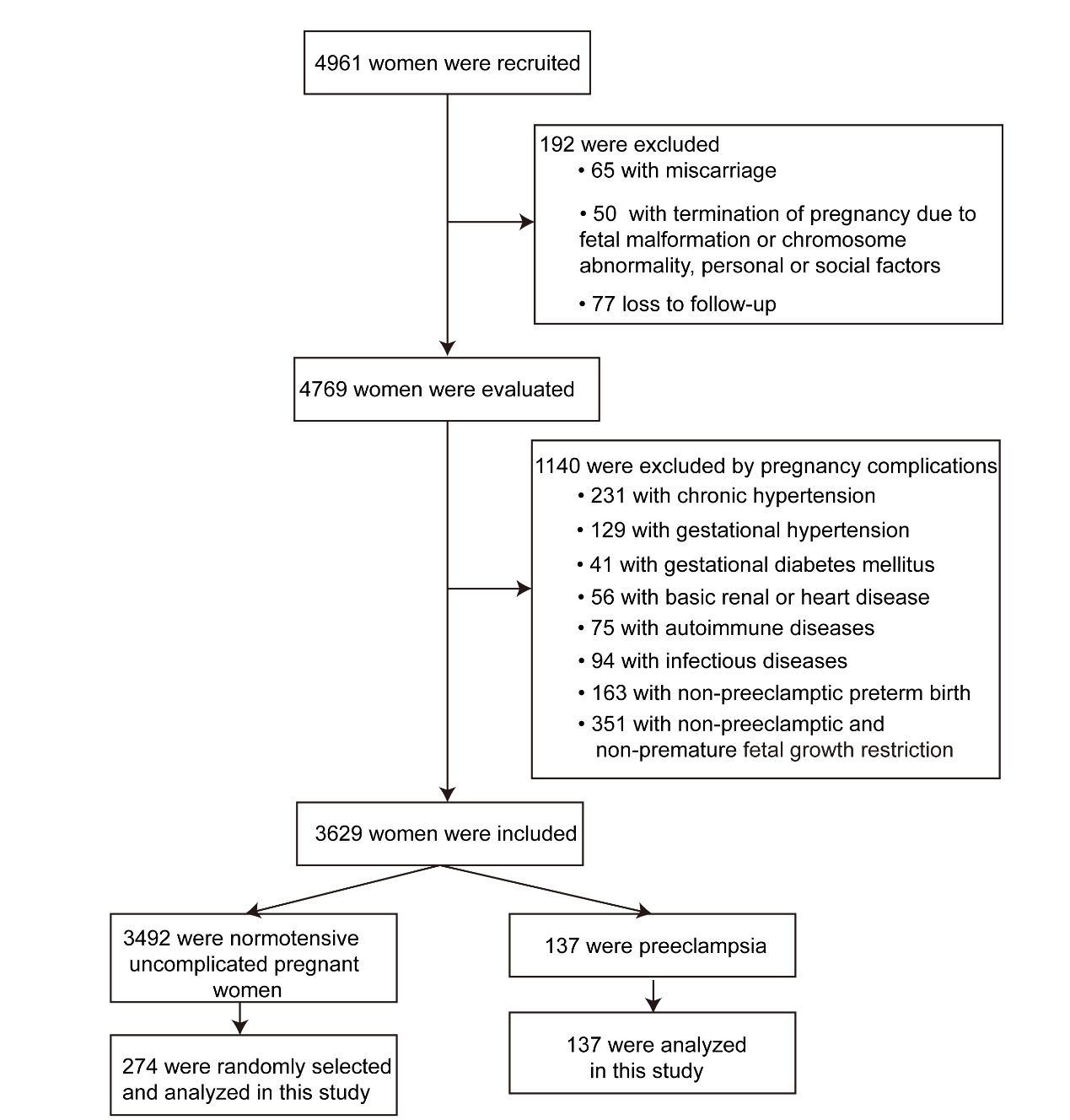

**Supplementary Figure 1. Flow diagram of participants in the longitudinal cohort.**

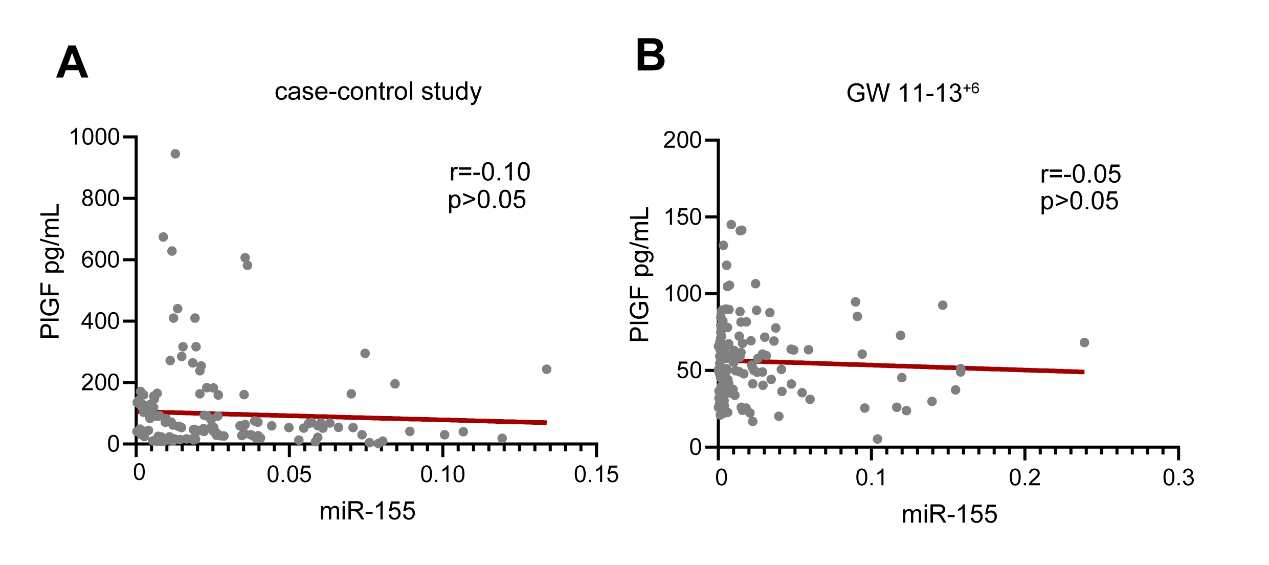

**Supplementary Figure 2. The correlations between miR-155 and PlGF in serum from preeclampsia patients.**

**A,** The correlation between miR-155 and PlGF in serum from preeclampsia patients in the case-control study. **B,** The correlation between miR-155 and PlGF in serum from preeclampsia patients at GW 11-13^+6^ in the longitudinal study.GW, gestational week.

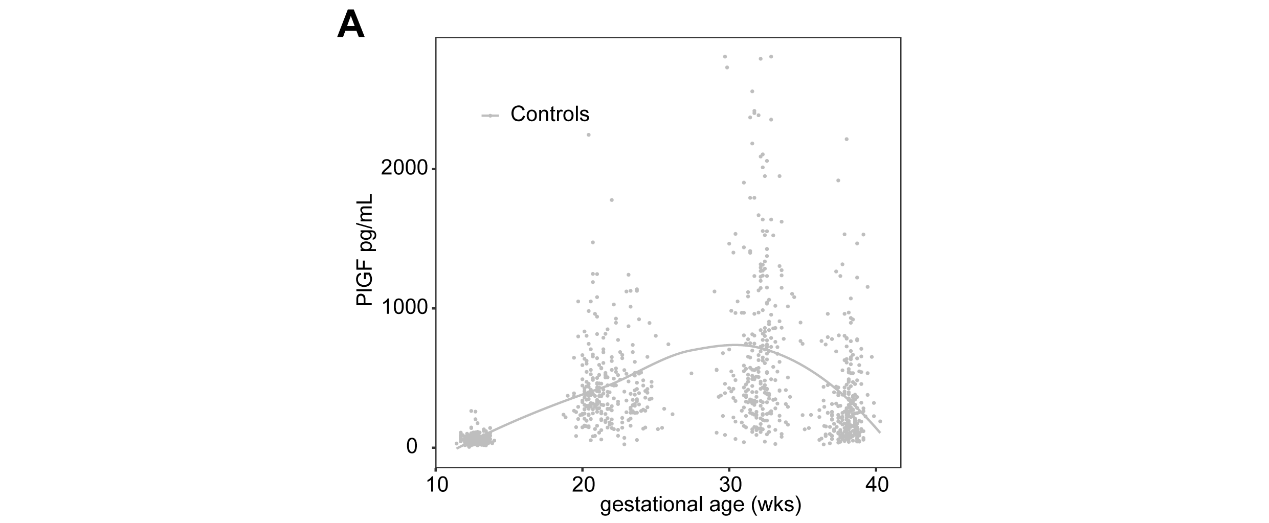

**Supplementary Figure 3. The PlGF concentration in the controls from longitudinal cohort study.**

**Supplementary Table 1. The number of serum samples taken in the longitudinal study.**

| Time | Controls | PE | Total |
| --- | --- | --- | --- |
| GW 11-13^+6^ | 274 | 137 | 411 |
| GW 19-23^+6^ | 274 | 137 | 411 |
| GW 30-33^+6^ | 274 | 121 | 395 |
| GW 35-38^+6^ | 274 | 72 | 346 |
| Total | 1096 | 467 | 1563 |

Serum samples were obtained from all preeclamptic patients from GW 11-13^+6^ to 19-23^+6^. Due to the childbirth, the number of serum samples decreased from GW 30-33^+6^ to 35-38^+6^. PE, preeclampsia; GW, gestational week

**Supplementary Table 2.** **The rate of cases with high miR-155 levels in the longitudinal study.**

| High miR-155 | GW 11-13^+6^ | GW 19-23^+6^ | GW 30-33^+6^ | GW 35-38^+6^ |
| --- | --- | --- | --- | --- |
| Controls, % (n/N) | 4.7 (13/274) | 4.7 (13/274) | 4.7 (13/274) | 4.7 (13/274) |
| PE, % (n/N) | 29.9 (41/137) | 27.7 (38/137) | 29.8 (36/121) | 30.6 (22/72) |

PE, preeclampsia; GW, gestational week. Data are expressed as percentages (n/N).

**Supplementary Table 3**. **The rate of cases with low PlGF levels in the longitudinal study.**

| Low PlGF | GW 11-13^+6^ | GW 19-23^+6^ | GW 30-33^+6^ | GW 35-38^+6^ |
| --- | --- | --- | --- | --- |
| Controls, % (n/N) | 4.7 (13/274) | 4.7 (13/274) | 4.7 (13/274) | 4.7 (13/274) |
| PE, % (n/N) | 13.1 (18/137) | 19.0 (26/137) | 24.0 (29/121) | 8.3 (6/72) |

PE, preeclampsia; GW, gestational week. Data are expressed as percentages (n/N).

**Supplementary Table 4.** **Clinical characteristics of normotensive controls with high miR-155 and normal miR-155.**

|  | Normotensive uncomplicated controls | |  |
| --- | --- | --- | --- |
| Analyzed items | Normal miR-155 | High miR-155 | *p*.value |
|  | n=594 | n=30 |  |
| Maternal age, y | 29.7 ± 2.8 | 29.9 ± 3.1 | >0.05 |
| Pre-pregnancy BMI, kg/m^2^ | 23.8 ± 2.5 | 24.2 ± 3.8 | >0.05 |
| GW at delievry, wks | 38.9 ± 5.3 | 37.3 ± 4.1 | <0.05 |
| IVF, n (%) | 78 (13.1) | 3 (10.0) | >0.05 |
| Multipara, n (%) | 85 (14.3) | 1 (3.3) | >0.05 |
| SBP, mmHg | 114.3 ± 9.5 | 120.4 ± 9.8 | <0.01 |
| DBP, mmHg | 72.4 ± 8.7 | 77.6 ± 11.6 | <0.05 |
| Birthweight, g | 3447 ± 586.3 | 3185 ± 603.2 | <0.05 |

BMI, body mass index; GW, gestational week; IVF, in-vitro fertilization; SBP, systolic blood pressure; DBP, diastolic blood pressure; The SBP and DBP were described as the highest blood pressure during admission.

Continuous variables are expressed as mean ± SD. Discrete variables are expressed as n (percentages).

**Supplementary Table 5. Clinical characteristics of normotensive controls with low PlGF and normal PlGF.**

|  | Normotensive uncomplicated controls | |  |
| --- | --- | --- | --- |
| Analyzed items | Normal PlGF | Low PlGF | *p*.value |
|  | n=594 | n=30 |  |
| Maternal age, y | 29.8 ± 3.2 | 29.6 ± 2.8 | >0.05 |
| Pre-pregnancy BMI, kg/m^2^ | 24.3 ± 2.7 | 23.7 ± 3.2 | >0.05 |
| GW at delievry, wks | 38.7 ± 4.5 | 37.6 ± 3.8 | <0.05 |
| IVF, n (%) | 85 (14.3) | 1 (3.3) | >0.05 |
| Multipara, n (%) | 84 (14.2) | 2 (6.7) | >0.05 |
| SBP, mmHg | 116.4 ± 8.3 | 115.7 ± 9.8 | >0.05 |
| DBP, mmHg | 73.1 ± 7.9 | 72.5 ± 8.6 | >0.05 |
| Birthweight, g | 3431 ± 560.4 | 3243 ± 586.3 | <0.05 |

BMI, body mass index; GW, gestational week; IVF, in-vitro fertilization; SBP, systolic blood pressure; DBP, diastolic blood pressure; PlGF, placental growth factor. The SBP and DBP were described as the highest blood pressure during admission.

Continuous variables are expressed as mean ± SD. Discrete variables are expressed as n (percentages).

**Supplementary Table 6.** **The medical history, clinical presentations and pregnant outcomes between patients with high miR-155 and high miR-155 accompanied with low PlGF.**

|  | PE | |  |
| --- | --- | --- | --- |
| Analyzed items | High miR-155 | High miR-155 with low PlGF | *p*.value |
|  | n=96 | n=30 |  |
| Maternal age, y | 30.3 ± 4.8 | 29.9 ± 5.8 | >0.05 |
| Pre-pregnancy BMI, kg/m^2^ | 24.1 ± 3.2 | 24.2 ± 3.5 | >0.05 |
| IVF, n (%) | 20 (20.8) | 4 (13.3) | >0.05 |
| Abortion history, n (%) | 31 (32.3) | 6 (20.0) | >0.05 |
| Multipara, n (%) | 15 (15.6) | 7 (23.3) | <0.01 |
| PE history, n (%) | 12 (80.0) | 5 (71.4) | <0.01 |
| Maternal assessment and diagnosis |  |  |  |
| SBP, mm Hg | 161.4 ± 16.6 | 164.3 ± 20.8 | >0.05 |
| DBP, mm Hg | 97.0 ± 12.5 | 101.4 ± 13.2 | >0.05 |
| SBP≥160 and/or DBP ≥110 mm Hg, n (%) | 61 (63.5) | 23 (76.7) | >0.05 |
| Proteinuria, mg/24h | 2851 ± 2471 | 3858 ± 2934 | >0.05 |
| Serum creatinine, μM | 50.5 ± 9.5 | 53.9 ± 11.3 | >0.05 |
| Serum uric acid, μM | 380.8 ± 88.4 | 385.9 ± 103.1 | >0.05 |
| AST, U/L | 31.1 ± 30.6 | 32.2 ± 19.5 | >0.05 |
| ALT, U/L | 23.8 ± 20.2 | 25.1 ± 18.8 | >0.05 |
| Platelet, ×10^9^/L | 159.2 ± 61.3 | 150.6 ± 66.7 | >0.05 |
| Platelet < 100×10^9^/L | 17 (17.7) | 4 (13.3) | >0.05 |
| Requiring urgent antihypertensive therapy (≥2 times), n (%) | 20 (20.8) | 8 (26.7) | >0.05 |
| HELLP, n (%) | 5 (5.2) | 1 (3.3) | >0.05 |
| Visual disturbance, n (%) | 15 (15.6) | 4 (13.3) | >0.05 |
| Hypertensive retinopathy, n (%) | 13 (13.5) | 6 (20.0) | >0.05 |
| Placental abruption, n (%) | 7 (7.3) | 2 (6.7) | >0.05 |
| MICU, n (%) | 3 (3.1) | 1 (3.3) | >0.05 |
| The early-onset PE, n (%) | 50 (52.1) | 15 (50.0) | >0.05 |
| Pregnancy outcome |  |  |  |
| GW at delivery, wks | 35.5 ± 3.6 | 33.8 ± 3.7 | <0.05 |
| Male infant, n (%) | 51 (53.1) | 16 (53.3) | >0.05 |
| Birthweight < 3th, n (%) | 21 (21.9) | 7 (23.3) | >0.05 |
| Apgar ≤ 7 at 1 min, n (%) | 6 (6.3) | 1 (3.3) | >0.05 |
| Apgar ≤ 7 at 5 min, n (%) | 3 (3.1) | 1 (3.3) | >0.05 |
| NICU, n (%) | 23 (24.0) | 8 (26.7) | >0.05 |

PE, preeclampsia; BMI, body mass index; IVF, in-vitro fertilization; SBP, systolic blood pressure; DBP, diastolic blood pressure; AST, aspartate aminotransferase ALT; alanine aminotransferase; MICU, maternal intensive care unit admission; GW, gestational week; NICU, neonatal intensive care unit admission; PlGF, placental growth factor. The SBP and DBP were described as the highest blood pressure during admission.

Continuous variables are expressed as mean ± SD. Discrete variables are expressed as n (percentages).
